# Jet Lag Does Not Impact Football Performance: Using Randomization Inference to Handle Complexity

**DOI:** 10.1101/2023.10.19.23296960

**Authors:** Matthew S. Tenan, Ali R. Rezai, Andrew D. Vigotsky

## Abstract

**Introduction:** It is commonly accepted that traveling across time zones affects sport performance (*i.e.*, via jet lag). This belief is based on poor quality evidence for team sports and simplistic analyses, such as *t*-tests and linear regression, to explore complex phenomena. For instance, Roy & Forest used such analyses to examine win percentages for the NFL, NBA, and NHL, concluding that East Coast teams were disadvantaged. Similarly, Smith et al. primarily used *t*-tests to show that West Coast NFL teams were more likely than East Coast teams to beat the Vegas spread in evening games (non-coastal teams were omitted). Neither analysis considered time zone change or game time as continuous constructs nor did they account for important contextual information. We used modern causal inference methods and a decade of collegiate football games to determine if jet lag and kickoff time have any causal effect on beating the Vegas spread. This required fitting nonlinear splines for both data re-weighting and analysis; however, using weights in a generalized additive model (GAM) presents challenges for standard frequentist inferences. Thus, non-parametric simulations were developed to obtain valid causal inferences via randomization inference (RI).

**Methods:** Pro Football Focus data from college football seasons 2013–2022 were paired with time zone data from Google Maps, weather data from gridMET, and Vegas spread data from collegefootballdata.com. GAM-based propensity scores were calculated from turf type, precipitation, humidity, temperature, and wind speed. These propensity scores orthogonalized the variables relationship to the treatments (*i.e.*, game time and hours gained due to time zone change), consistent with the Potential Outcomes framework. The propensity scores were used to weight the observations in a GAM logistic regression, which modeled beating the Vegas spread as a function of a splined effect modification for game time and hours gained in travel. Since valid standard errors cannot be calculated from GAMs with weights, we used RI to compare the effect modification to a null model. We simulated 5,000 datasets of random treatments under the positivity assumption. Each RI dataset was analyzed with the same GAM used for the observed data to obtain a distribution of noise *F*-statistics. The real data *F*-statistic was contrasted to the RI distribution for inferences.

**Results:** The real data were compatible with the null hypothesis of no effect for hours lost/gained in travel and game time (*P* = 0.142).

**Conclusion:** We need to rigorously interrogate assumptions regarding what affects performance in team sports. There is no clear indication that jet lag and game time affect team performance when appropriate analyses are performed in a causal inference framework. Similarly, rigorous analysis should be undertaken to confirm or refute other assumptions in sport science, such as workload management, sleep practices, and dietary/supplementation regimens.

## 1. Introduction

It is commonly accepted that traveling across time zones affects sports performance via “jet lag”.^1–7^ This concept has likely gained broad acceptance because non-athletes often traverse multiple time zones and experience a perception that “something is off” or have irregular sleep patterns, so it is straightforward for non-athletes to accept the premise that jet lag affects sports performance with minimal resistance. One complication is distinguishing travel fatigue from jet lag.^6^ Travel fatigue is a non-medical condition resulting from frequent travel with minimal recovery, resulting in persistent fatigue, generalized recurrent illness, and potential mood changes.^8^ Jet lag is a medical condition and specific to the travel direction (east→west/west→east) and the number of time zones crossed, resulting in distinctive sleep disturbances, daytime fatigue/sleepiness, and impaired mental/physical concentration.^9,10^ Whereas it seems apparent that travel fatigue can be mitigated by optimizing travel logistics, such as limiting connecting flights or providing appropriate recovery between travel, jet lag is less forgiving: If an athlete must travel from one location to the next, it requires a specific direction and crossing an exact number of time zones. Despite the conceptual overlap between travel fatigue and jet lag, they have different cause-effect structures and thus provide different opportunities for intervention.

Domestic travel, where flights traverse three or fewer time zones, provides the most common opportunity to experience jet lag in team sports. Even individual sports, such as track and field or swimming, will have many domestic meets punctuated by a large-scale international meet. The lower volume of international meets, which necessitate traveling more than three time zones, make it challenging to statistically assess (low number of samples) and easier to develop adaptations for the athlete (*e.g.*, arrive earlier to allow adaptation and recovery). As such, most published research has focused on how domestic jet lag may impact individual human performance and team performance.

### 1.1. Current Evidence on Jet Lag & Sport Performance

There is minimal and low-quality evidence that jet lag impacts an individual athlete’s physical or cognitive capabilities. The highest quality study used a randomized cross-over design of 10 athletes in control (no flight) and a simulated 5-hour domestic flight, showing that jet lag had no statistically significant effects on jump height, jump power, jump peak velocity, intermittent sprint performance, sleep duration, sleep latency, sleep efficiency, number of awakenings, duration of awakenings, various perceptual measures (fatigue, soreness, anger, confusion, vigor, and depression), or salivary cortisol, though blood oxygen saturation was lower following travel.^11^ A small cohort of professional soccer players (n=6) showed no statistically significant effect on sleep patterns or perceptual measures of sleepiness and recovery after repeated 5-hour air travel across two time zones,^12^ though such a small study may be underpowered. A somewhat larger study of 19 Australian rules footballers indicated that traveling 1–2 time zones resulted in no measurable change in objective sleep metrics, but a small perceptual decrease in sleep quality.^13^ Both the small soccer cohort and the Australian rules footballer study noted minor differences in in-match metrics when comparing home and away games, but it is impossible to attribute this difference to the travel itself.^12,13^

In contrast to individual-level studies examining jet lag and human performance, ‘big data’ studies generally show an apparent effect of jet lag decreasing team performance.^14–19^ However, all of these studies employ univariate analyses^16–19^ and/or infer causality from multivariable correlations.^14,15,18,19^ Such modeling is not strongly informed by theory (*i.e.*, a putative causal structure) and may thus lead to fallacious causal inferences. For example, three studies only include data for teams originating and playing in the Pacific and Eastern time zones,^16,18,19^ omitting major sports markets and failing to effectively capture anything resembling real-world dynamics. Given the noted analytical issues, it should not be surprising that studies have paradoxically shown unidirectional jet lag-based decrements in performance for both westward travel (basketball^16,17^, hockey^17^) and eastward travel (football^18^, baseball^14,19^, basketball^15^). Most studies examining overall game outcomes have also indicated that this effect only occurs in evening games.^17–19^ The lack of “null” findings may be attributable to positive publication bias,^20^ but how does one explain apparent effects in different travel directions between sports? *Post hoc* justifications about commonalities between basketball and hockey (where westward travel is problematic) and associated differences with football, baseball, and basketball (where eastward travel is problematic) are common but unprincipled. Moreover, modeling missteps preclude causal inferences. For instance, team-level studies have not treated game time or jet lag as the continuous or interval variables that they are in the real world, nor did they approach their analyses with an eye toward causation. Only one study attempted to account for a team’s expectation of winning a game by benchmarking their performance against the Vegas spread.^18^ The ‘wisdom of the crowd’ Vegas spread, while imperfect, appears to be the most reasonable method of determining if a team under- or over-performs expectations.

### 1.2. Causal Inference for Complex Data Structures

All models are, by design, simplifications of a complex reality. The estimand of a causal inference problem is the true effect of an intervention—what the study aspires to capture.^21^ As the estimand is a simplified theoretical model, even complex models can often be mathematically described with relative ease. For example, the estimand equation for the directed acyclic graph in Figure 1 (Main Analysis) could be written as:

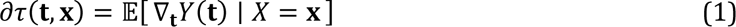

**Figure 1.**
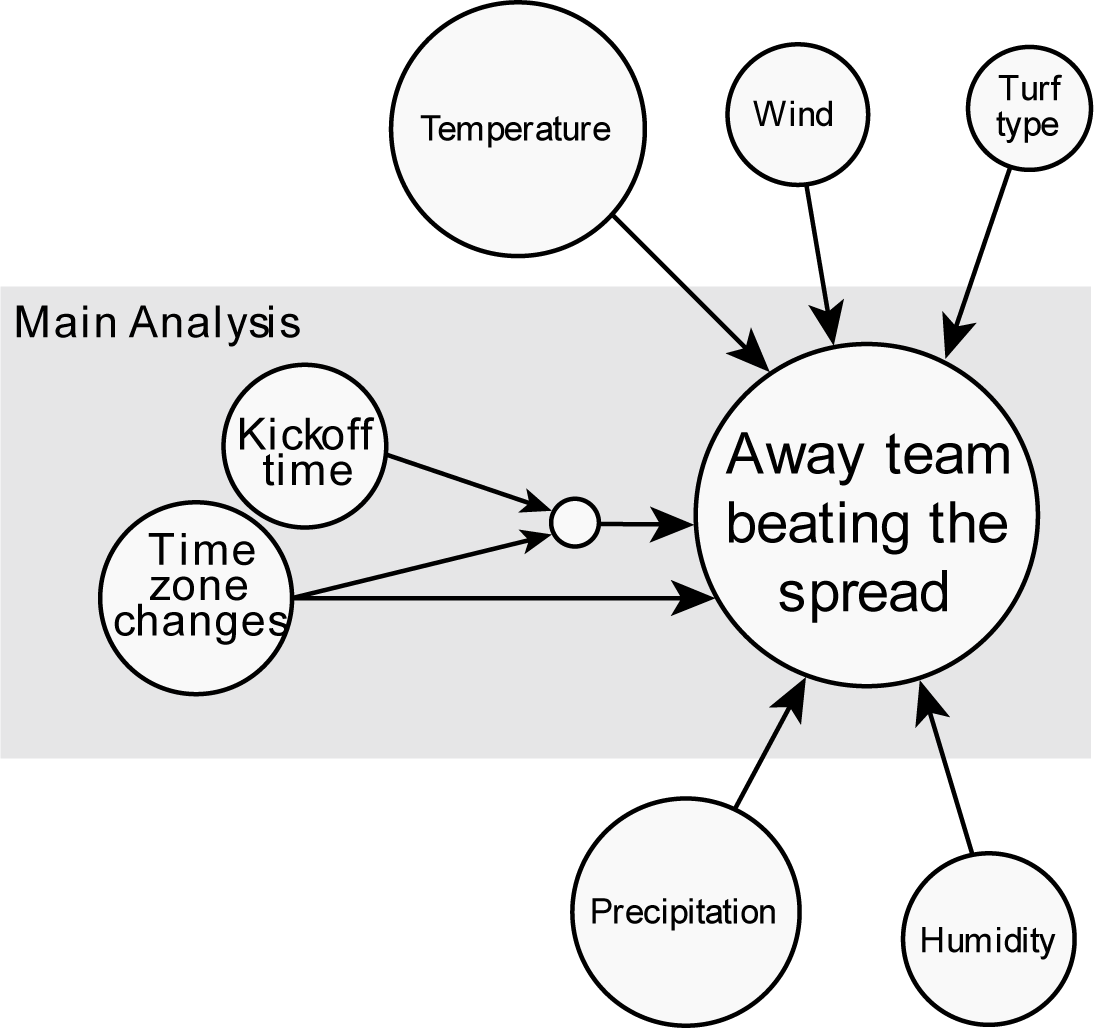
DAG depicting the theoretical causal structure.

In the current study, we will test the above estimand equation (1), which is the marginal conditional average treatment effect (CATE). This is the instantaneous rate of change (∂) of the average treatment effect (*τ*) for each hour gained/lost in travel (**t**), conditional on kickoff time (**x**). This is estimated from the expected value (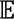) of the partial derivative (∇**_t_**) of the away team’s performance (Y(·)) as a function of hours gained/lost in travel on that game’s performance (Y(**t**)), conditional on the kickoff time for that game (X). This is different than a conceptually more straightforward but less omnibus examination of two counterfactuals (*e.g.*, the difference between losing three hours in travel and gaining three hours in travel, conditional on kickoff time), which artificially dichotomizes the “hours gained/lost” dynamic into a binary question. While equation 1 is elegant, how one arrives at the real-world estimate of the estimand based on real-world data does often not cleanly conform to an estimand equation.

When confronted with complex data structures that do not cleanly conform to a directed acyclic graph or estimand equation, one common approach is to claim that this data structure itself is “non-causal” in nature.^22^ However, ignoring complex data structures has been shown to create a mismatch between a model estimate and desired estimand and, as a result, inflate error rates for inferences.^23–25^ For this reason, the potential outcomes framework^26^ is often preferred over the structural causal model^27^ when the real-world data has a complex or nested underlying structure. The strength of the potential outcomes framework is that it is grounded in approximating a randomized control trial and thus has a multitude of statistical procedures and methods from which to analyze complicated data often seen in sport.

### 1.3. Randomization Inference – Sometimes the Only Valid Solution

When an intervention is dichotomous (*e.g.*, treatment and control) or polychotomous (*i.e*., different treatments), there are well-established group-based matching and analysis methods for causal inference.^28^ Most ‘off-the-shelf’ causal inference toolboxes assume that data are independent and identically distributed (i.i.d.) random variables.^29^ When an intervention is continuous in nature and data have a known hierarchical structure (*i.e.*, are not i.i.d.), which is common in sport, analyses based on inverse probability weights (IPW) are the only current option available.

Although various IPW methods are firmly established for linear versions of clustered or time-series data,^30,31^ none can handle penalized splines to account for nonlinear relationships, commonly called generalized additive models (GAMs) and their extension, generalized additive mixed models. To our knowledge, only two popular software packages implement weighting in GAMs: STATA and R.^32,33^ Both software packages implement frequency weights, which will produce correct parameter estimates but invalid standard errors, precluding valid interval coverage and frequentist hypothesis testing rates. Frequency weights are predefined counts, which assume each observation *y_i_* can be counted *w_i_* times. In contrast, IPW assumes that *y_i_*was observed with a probability 1/*w_i_*, altering the weights to represent the target population based on empirical estimates from the propensity model. These assumptions lead to disparate standard error estimators. Since frequency weights are predefined and are akin to duplicating rows in a data frame, their standard errors are well-defined. However, IPWs are *estimated* rather than defined or assumed, meaning that IPWs themselves have uncertainty and structure that classical estimators do not consider. Standard errors that consider the unique properties of IPWs have not yet been developed for GAMs.

In many scenarios, including linear regression, bootstrapping would be a valid solution for extracting appropriate standard errors and confidence intervals; however, it has been previously demonstrated that bootstrapping is not point-wise valid with GAMs and could lead to misleading inferences,^34,35^ a concept that seems to have been overlooked in both the epidemiology^36^ and political science^37^ literature. GAMs have the added benefit of being able to model a treatment effect that is a nonlinear interaction between continuous variables or a mix of multiple categorical and continuous variables. This is an important flexibility in sport where athletes may simultaneously receive many “treatments” that may affect a potential outcome. After an extensive survey of the literature and computational software, we concluded that short of deriving new estimators, randomization inference was the only viable solution to our current question: Does jet lag, conditional on kickoff time, have a causal effect on whether a team beats the Vegas spread (*i.e.*, performs relative to expectations)?

Randomization inference assesses all (or a sufficiently large sample) of the possible treatments that could have been assigned to a unit (team). The observed outcome is then compared to the distribution of randomized potential outcomes.^38^ While much hypothesis testing relies on theoretical distributions (*e.g.*, *t*-tests = *t*-distribution; analysis of variance = *F*-distribution, *etc*.), randomization inference fully describes the sampling distribution under the null hypothesis and does not rely on an approximation of a theoretical distribution. The specific metric or test statistic used to build the null distribution and draw inferences depends on the research question and analysis. For example, many studies have evaluated the model coefficient for the factor of interest. In contrast, if a system of multiple coefficients is of interest, *F*-statistics are a more natural choice, and a researcher can compare the treatment data’s *F*-statistic to the corresponding null model of potential outcomes’ *F*-statistics.^38^ Under the potential outcomes framework, randomization inference is a flexible analytic solution that works under all circumstances with minimal assumptions; thus, it can be leveraged to determine how extreme an observed outcome is relative to a noise model, so long as the scientist/analyst can conceptualize and simulate all possible treatments.^39^

## 2. Methods

The data for this work arose from multiple sources: Pro Football Focus (PFF), Wikipedia, Google Maps, gridMET, collegefootballdata.com, and the ‘lutz’ R package.^40^ Across the sources, team names, university names, or geographic locations required minor alterations so that the fuzzy matching procedure^41^ rendered an accurate join. First, the NCAA Football teams in PFF were matched with the physical locations of each institution from Wikipedia.^42^ The location of each institution was then converted into a latitude and longitude via the Google Maps API and used to obtain the time zone of each location via the ‘lutz’ R package. The PFF API was then queried for all game information from the 2013–2022 NCAA football seasons. The PFF game information included the stadium at which the game was played and the latitude/longitude coordinates, which were subsequently cross-indexed to obtain the time zone for each stadium. During this process, it was recognized that some stadium longitudes were incorrect in the PFF database, which was manually fixed and PFF was notified of the issue. The corrected stadium latitude, longitude, and game date were then matched with associated meteorological data from the gridMET database.^43^ Finally, each game was matched with the historical consensus Vegas spread data from collegefootballdata.com via team names and game scores for each season.^44^ This process rendered 6,245 complete games for analysis. Throughout this process of data aggregation, accuracy was continually manually assessed.

All game times were standardized to the United States Eastern Time Zone and rounded to their nearest hour on a 24-hour clock. The time difference between the away team’s time zone and the time zone in which the game was played was calculated. Any observations where the home team was playing at a stadium located in a different time zone than their institution were removed. All games where either the stadium’s location or the away team’s institutional location was not in the continental United States were removed. As weather data is not pertinent when games are played inside, all weather variables for inside games were set to ‘0’. Finally, a binary variable was created indicating if the away team beat the Vegas spread where ‘1’ indicates beating the spread and ‘0’ indicates not beating the spread in a particular game.

### 2.1. Causal Structure

The outcome variable of interest, the away team beating the Vegas spread, encodes a lot of underlying information, such as player personnel (including known injuries to important players), coaching, “home field advantage”, and other expectations about team play and capabilities. One thing the pre-game consensus Vegas spread cannot fully account for is weather patterns that may impact gameplay during the game (Figure 1). Additionally, our DAG assumes that the Vegas spread is not capturing causal information regarding hours gained/lost from time zone changes, the conditional effect of kickoff time on time zone change, and turf type (field turf, artificial turf, or real grass). The effect modification of kickoff time is shown on the DAG in accordance with the conventions proposed by Attia et al.^45^ Because the weather variables (highest temperature, highest wind speed, highest humidity, and precipitation) and turf type have causal paths to the outcome only, they should be included in the propensity score under the potential outcomes framework, and including these variables increases the precision of the estimated effect without impacting bias.^46^

### 2.2. Propensity Score Development & Validation

A GAM was used to create the propensity score to account for nonlinearities as well as incorporate an assumed multi-level structure, which was hypothesized to be inherent in the data. In the context of the current experiment, we want it equally likely that each game could have theoretically been “assigned” to any level of the effect modification “treatment” (kickoff time and hours gained/lost from time zone changes, see Figure 1).^47^ For the purposes of propensity score development, a multivariate generalized additive mixed model modeled the multivariate Gaussian where games are nested within away teams. This structure makes theoretical sense as a team like the University of North Carolina-Chapel Hill on the East Coast will have time zone changes ranging from 0 to +3 as they travel from east to west, and a team such as the University of Texas-Austin will have time zone changes ranging from −1 to +2, depending on if they travel to the east coast or towards the west coast. Similarly, many East Coast teams are likely to have a higher probability of playing earlier in absolute time than West Coast teams as they will play relatively more games in the Eastern Time Zone (*e.g.*, the University of Southern California would not play a home game at noon Eastern Time, as this is 9 am Pacific Time). The necessity of the multilevel propensity score for both time zone changes and kickoff time was confirmed by nonnegligible away-team intraclass correlation coefficients (ICC) of 0.29 and 0.16,^48,49^ respectively.

All weather predictors were continuous variables and allowed to vary nonlinearly using penalized thin plate regression splines. The propensity scores were then cluster-mean stabilized as previously described.^30^ After stabilization, the covariate balance of the propensity score was checked to verify that each factor was balanced across the effect modification treatment.^50^ The vast majority of the literature on IPW validation of covariate balance only assesses univariate treatments and not a joint effect modification treatment.^51^ The most commonly used metric to assess covariate balance for continuous data, the Pearson correlation coefficient, was inappropriate in our case. Thus, we took a two-pronged approach to determine if our IPW was effective in orthogonalizing the covariates to the effect modification treatment: 1) show that the IPW decreases the overall fit of the treatment to the covariate via increasing Akaike’s information criterion (AIC) and 2) examine the proportion of variance explained between the effect modification treatment and the individual covariates via the marginal *R*^2^ criterion.^52^ For the continuous covariates, this was assessed via linear multilevel models with observations clustered at the away team level (*e.g.*, precipitation was regressed on hours gained or lost in travel and (kickoff time × hours gained or lost in travel)). The balance assessment for grass type was slightly more complicated as a polychotomous covariate. In this case, we extracted the AIC from an analogous multinomial multilevel model with restricted maximal likelihood estimation, where we were unable to obtain an *R*^2^, and re-fit with a multinomial multilevel model with penalized quasi-likelihood estimation to obtain an *R*^2^. The goal of the balance diagnostics was to see the AIC reliably increase with the IPW and for *R*^2^ to be as low as possible, and certainly below a 0.1 threshold.^52,53^ The adjusted and unadjusted *R*^2^ variance explained statistics for the effect modification treatment and the covariates can be seen in Figure 2 and the AIC metrics are available in Table 1.

**Figure 2.**
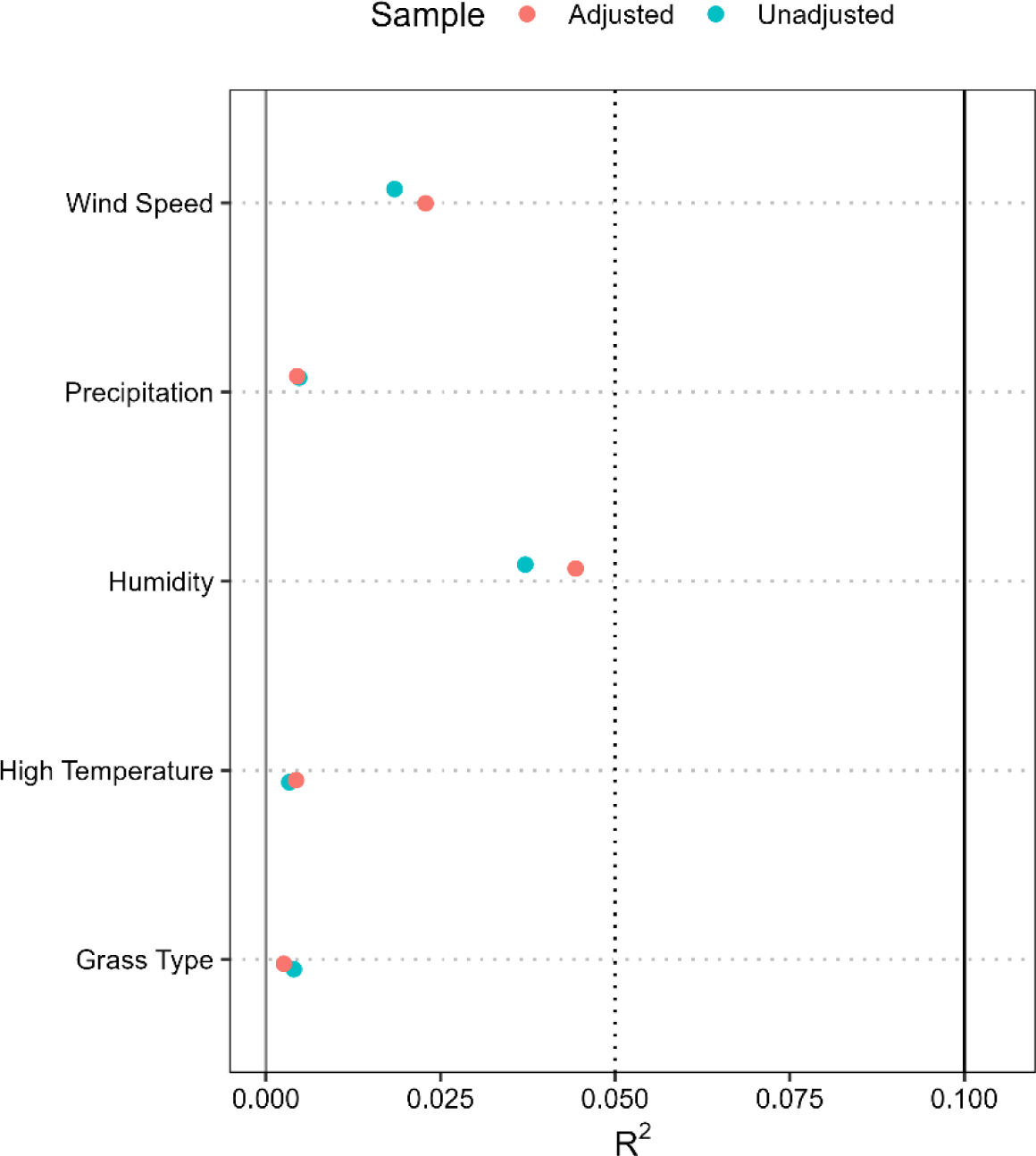
R^2^ statistics for the unweighted (unadjusted) and weighted (adjusted) proportion of variance assessments between the effect modification treatment and the covariate of interest. The solid vertical line at 0.100 is a commonly applied threshold and the dashed line at 0.050 is the desired threshold we applied.

**Table 1.** IPW-adjuscted models increase the AIC for each covariate, indicating a lower association between the covariate and the effect-mediated treatment.

| Covariate | Unadjusted AIC | Adjusted AIC | Weighting Differential |
| --- | --- | --- | --- |
| Precipitation | 5,833 | 6,179 | +346 |
| Humidity | 55,495 | 55,720 | +225 |
| Temperature | 54,701 | 54,946 | +245 |
| Wind Speed | 34,742 | 34,820 | +78 |
| Grass Type | 2,682 | 2,749 | +67 |

### 2.3. Causal Inference via Randomization Inference

In a GAM logistic regression with propensity score weights, we can orthogonalize the secondary variables affecting the away team’s probability of beating the spread, enabling us to focus on the main problem of interest: “Does the gain or loss of hours due to travel, in combination with kickoff time, causally affect the probability of beating the Vegas spread?” To do this, we used a smoothed effect for hours gained/lost with 5-dimension bases and a tensor product interaction smooth for the effects of hours gained/lost and kickoff time with 7-dimension bases. We originally assumed that this model would require a nested structure, similar to that of the propensity score, but away team-level ICCs were negligible (0.015),^54^ suggesting this additional complexity, which introduced considerable computational demand, was unnecessary.

To our knowledge, there is no analytical software implementing probability sampling weights within a GAM framework. The weighting type implemented for GAMs in R and STATA is frequency weights (no weighting is offered for GAMs in SAS or Python), which appropriately model parameter estimates but not standard errors.^55^ Thus, we employed randomization inference of *F*-statistics to facilitate causal inferences.^38^ This involved fitting an intercept-only ‘null model’ to predict the outcome measure (away team beating the spread) while including the propensity score weights.

We then performed an analysis of variance (ANOVA) by comparing the null model with the full model (including the smoothed effects) to obtain an *F*-statistic. While this *F*-statistic is not valid for inference based on a standard *F-*distribution, it can be contrasted to a distribution of *F*-statistics generated from all potential outcomes (or a sufficiently large sample of potential outcomes), to determine how a model fit using our real data compares to a model fit using randomly generated data (*i.e.*, randomization inference).^38^

To perform randomization inference, we need to build many datasets of treatments (*i.e.*, noise treatments) while maintaining the current positivity status. Positivity is the concept that all games should be equally exposed to each given treatment—something that our study, by definition, violates (termed structural or deterministic positivity).^56^ Section 2.2 noted the clustered nature of time zone changes and kickoff times; this same dynamic plays out with the randomization inference datasets. Our noise treatments should be theoretically possible and not suggest that the North Carolina Tar Heels can play an away game where they lose two hours due to travel, which would imply they were playing somewhere in the Atlantic Ocean. The kickoff time positivity requirement is neither fully random nor fully structural. A school’s local time, location of their conference (e.g. Atlantic Coast Conference, Southeastern Conference, etc.), and the school makeup of that conference naturally create a differential probability of kickoff times (in the Eastern Time Zone) that a team is likely to play; however, given the substantial conference re-alignments occurring at present, it is unclear if the randomization inference noise data should adhere to previous proportions of kickoff times for each team or a fully random assignment. As such, we created both datasets to see if this affects our final inferences.

Five thousand noise treatment datasets, each consisting of 6,245 observations, were simulated. These 5,000 datasets were analyzed with the same GAM model used with the real data, and then an ANOVA was performed between that full noise model and the null noise model to render a “noise *F*-statistic”. The 5,000 noise *F*-statistics are then plotted in a histogram with the observed *F*-statistic. This distribution is then used to determine the compatibility of the observed *F*-statistic with the distribution of noise *F*-statistics *—i.e*., a *P*-value. As a matter of intellectual curiosity, the *P*-values from the naïve non-causal (*i.e.*, unweighted), naïve causal (*i.e.*, weighted), causal randomization inference with fully randomized kickoffs, and causal randomization inference with proportionally randomized kickoffs are reported to illustrate the dynamics of how inferences might change in these different scenarios.

## 3. Results

The resulting *P*-values for planned tests can be seen in Table 2. The randomization inference analyses— the only completely valid causal inference analyses—are both highly compatible with the null hypothesis that time zone changes, with or without an effect mediation of kickoff time, do not impact the probability of beating the Vegas spread. The non-causal analysis, simply assessing the non-linear relationship between effect-mediated travel time changes and the Vegas spread, was also compatible with the null hypothesis. As expected, inappropriately using frequency weights and an invalid hypothesis test (*i.e.*, naïve causal analysis) results in an overly liberal *P*-value and likely incorrect causal inference.

**Table 2.** Final *P*-values from the naïve relational analysis, naïve causal analysis with known errors, and the two causal randomization inference analyses.

|  | *P*-Value |
| --- | --- |
| Naïve Non-Causal Analysis | 0.387 |
| Naïve Causal Analysis | 0.028 |
| Randomization Inference w/Fully Random Kickoff Time | 0.133 |
| Randomization Inference w/Proportionally Random Kickoff Time | 0.142 |

There is no statistically significant causal effect of jet lag and kickoff time on the probability of the away team beating the Vegas spread. The ‘real data’ *F*-statistics can be contrasted against a large sample of the potential outcome *F*-statistics, both when the kickoff time is fully randomized and when it is kept proportional to historical data (Figures 3 and 4, respectively). The surface plot of point estimates for the observed data is depicted in Figure 5—point estimate probabilities are denoted by the black lines. The left-hand side of the surface plot illustrates eastward-traveling west coast teams (*i.e.*, ‘losing hours’ in time zone changes) and the right-hand side of the plot represents westward-traveling east coast teams (*i.e.*, ‘gaining hours’ due to changing time zones). All kickoff times have been standardized to the Eastern Time Zone; as such, the bottom-right quadrant of the surface plot is uncommon in the real world, as it implies an East Coast team traveling to the West Coast and playing between 9:00 am and 11:00 am Pacific Time.

**Figure 3.**
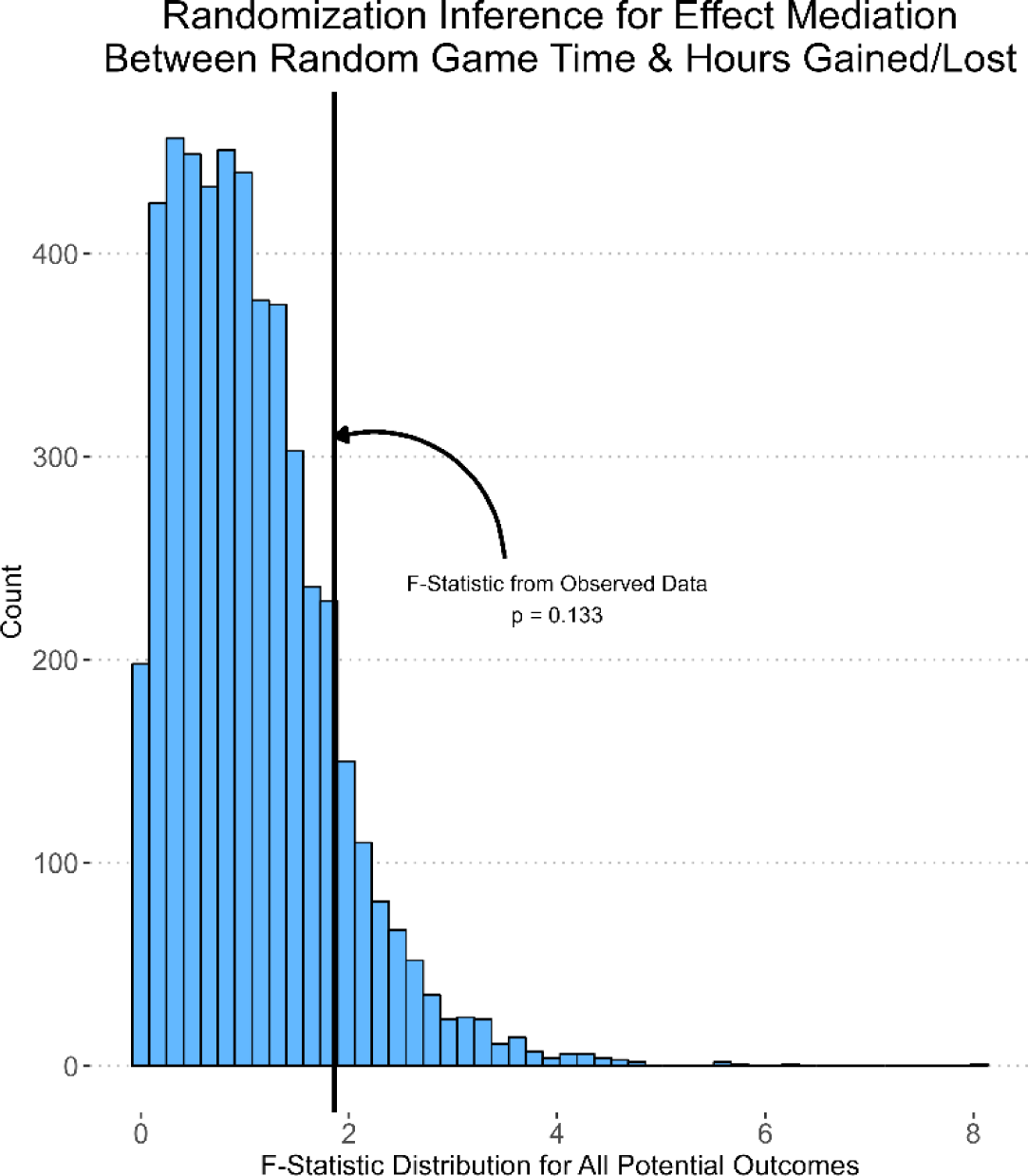
*F*-statistic for observed effects of jet lag and kickoff time contrasted against the distribution of *F*-statistics for 5,000 potential outcomes when kickoff time is fully randomized and hours gained/lost is fully random but consistent with positivity assumption.

**Figure 4.**
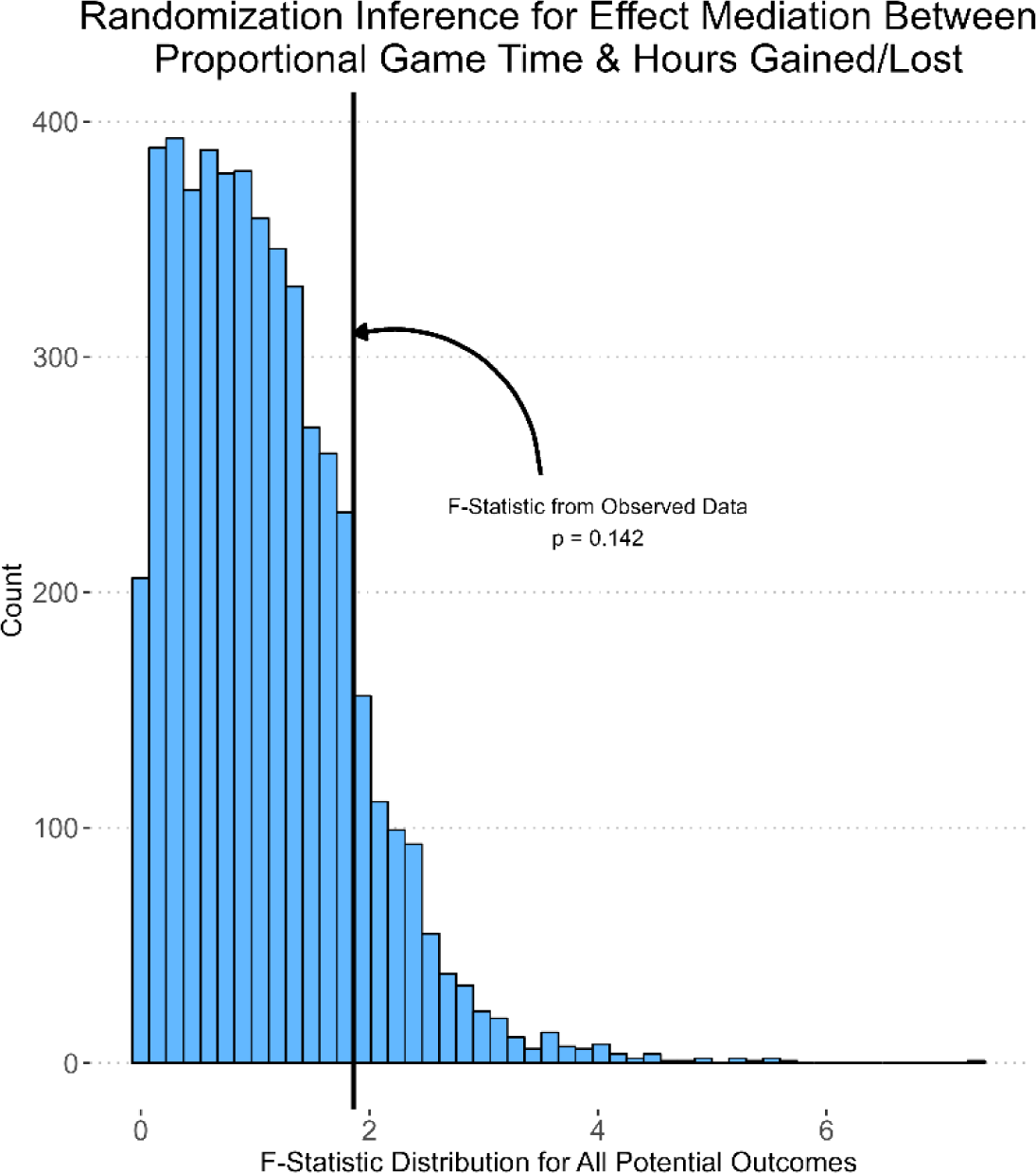
*F*-statistic for observed effects of jet lag and kickoff time contrasted against the distribution of *F*-statistics for 5,000 potential outcomes when kickoff time is randomized proportionally to existing data and hours gained/lost is fully random but consistent with positivity assumption.

## 4. Discussion

Using rigorous causal inference methods, we have shown there to be no discernable causal effect of jet lag and kickoff time on collegiate football team performance in the continental United States. Moreover, our jet lag data is likely contaminated with ‘information’ from travel fatigue that we are unable to remove with any available data. To remove any effects of travel fatigue, we would have needed data related to the away team’s travel itinerary and plans, which were not available for the representative subset of the 6,245 games examined. We can either assume that travel fatigue *does* have a causal effect on decreasing a team’s probability of beating the Vegas spread, in which case our null findings represent a liberal estimate of a jet lag × kickoff time effect (*i.e.*, the real effect is even lower than our point estimates) or travel fatigue *does not* have a causal effect on a team’s probability of beating the Vegas spread (*i.e.*, this travel fatigue effect either does not exist or is contained within the Vegas spread), in which case there should be no change in our estimates or inferential results. In either case, we can conclude that in a collegiate football population, there is no CATE of jet lag and kickoff time on beating the Vegas Spread (*i.e.*, team expected performance).

Previous studies showing conflicting effects of jet lag (with performance decrements in different geographic directions) and game time were all completed using professional team data.^14–19^ It is reasonable to assume that professional sports have more resources to anticipate, mitigate, and handle any potential effects of jet lag or travel fatigue. Furthermore, it is not unreasonable to imagine a selection bias where athletes more capable of rapidly adapting to travel perturbations (either from jet lag or fatigue) would be selected to move from the collegiate to the professional level. In other words, advancing from high school to university and university to professional levels necessitates more long-distance travel while continuing to perform at a higher level of play. Therefore, if we cannot discern an effect of jet lag and game time on a large dataset of collegiate football games, it certainly calls into question previous studies suggesting these effects existed in NFL teams,^17,18^ if not other professional sports.^14–17,19^ The surface plot of point estimates (Figure 5) further supports the idea of an unlikely effect from jet lag as they seem to not follow any sort of rationale or logic. Figure 5 could be interpreted to say that West Coast teams playing on the East Coast overperform expectations when games are played between noon and 5 pm local time and underperform expectations when games are played between 8 pm and midnight local time; East Coast teams playing on the West Coast overperform expectations when games are played between 11 am to 2 pm local time. In other words, the more time zones a team travels—whether eastward or westward—the more likely they are to (a) win if the game is early or (b) lose if the game is late. A large set of data across nearly a decade, high compatibility with the null hypothesis, and point estimates that defy explanation all indicate that no effect of jet lag on team performance is the most appropriate interpretation.

**Figure 5.**
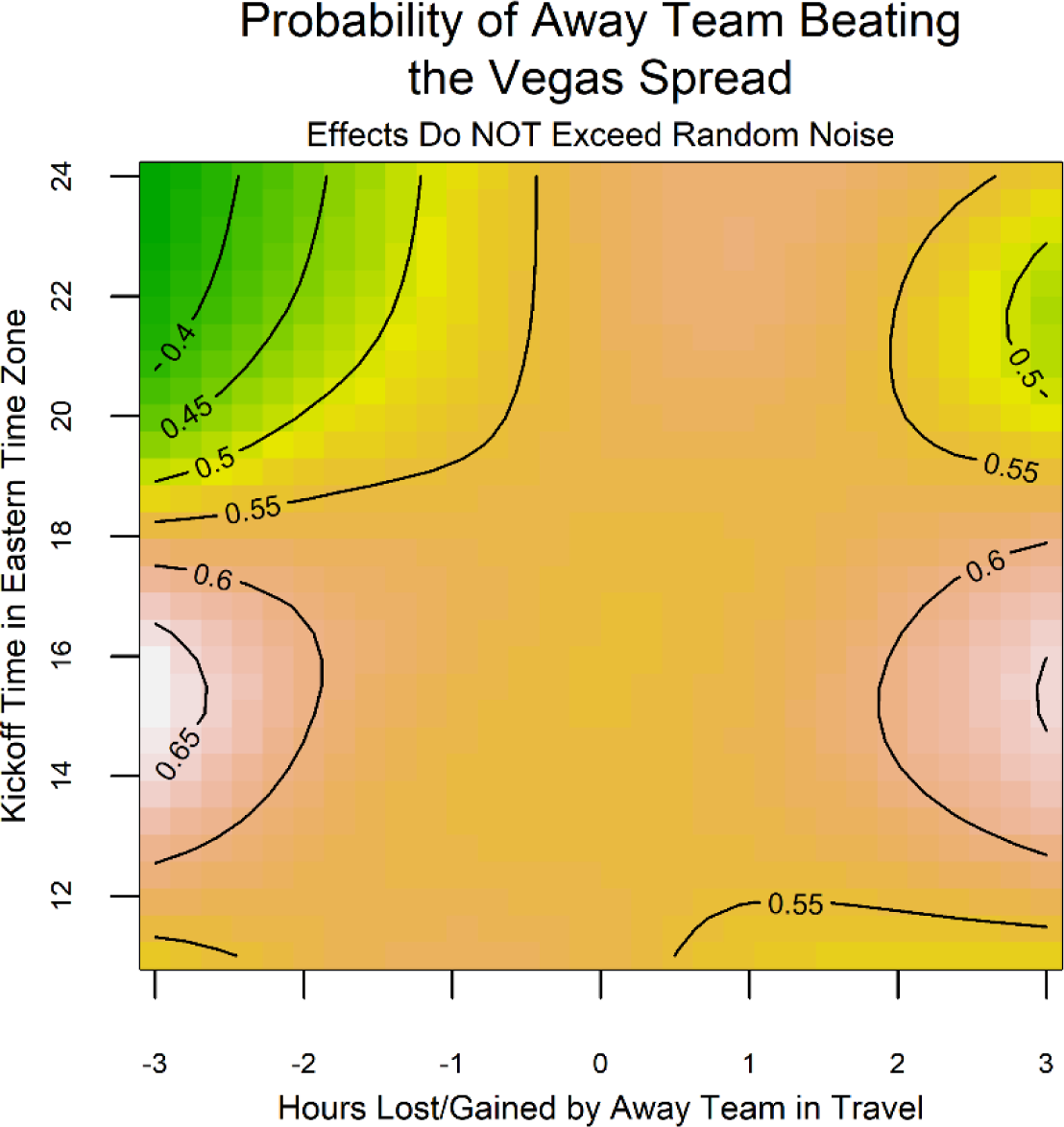
Surface plot of observed point estimates.

We believe Table 2 illustrates how a non-causal ‘relational’ analysis can sometimes reach more accurate inferences than a poorly conducted causal inference analysis. With the increasing number of widely accessible causal inference libraries available in the Python and R programming languages, it is likely that many untrained data scientists or statisticians are employing causal inference methods incorrectly, leading to poor inferences delivered to stakeholders.^57^ It is critically important for upper management within a sport organization to hire personnel with verifiable analytical training and skill sets for their desired tasks. Relatedly, it is critical for these organizations to have external consultants available to evaluate their internal analyses if they want to ensure that quality work is continually performed which increases team and organizational performance both from a sport and business perspective.

Complex questions require complex models that do not have a “textbook” solution from which to draw inferences and conclusions. For many complex models, bootstrapping is a general solution from which one can derive standard errors and confidence intervals.^58^ However, the bootstrap is not valid with penalized splines, and the typical solution, resampling the posterior, is not appropriate when using weights.^34,35^ As such, we propose randomization inference as an even broader solution than bootstrapping when it is necessary to make as few distributional or structural assumptions as possible for an inference problem. The primary requirement for randomization inference is that the general structure of the potential outcomes distribution must be able to be simulated.

Randomization inference is closely related to permutation testing. Indeed, it has been stated that these procedures are equivalent,^59^ randomization inference is a special case of permutation testing,^60^ and permutation testing is a special case of randomization inference.^61^ We agree with the work of Zhang & Zhao^39^ who argue that they are not equivalent methods and one does not subsume the other. Rather, randomization inference is based on randomized experimental design, while permutation testing is based on random sampling after data is collected under the required assumption of exchangeability.^62^ Specific to the current analysis, permutation testing could have been a viable solution had the permutations been clustered within the away team (failure to cluster permutations would violate the positivity requirement). However, this is probably not the best solution for two primary reasons: 1) much of the potential outcomes framework is based on the design of experiments and examination of all potential outcomes, so randomization inference is theoretically consistent whereas permutation testing is not, and 2) while nine years of game data is substantial, there is no theoretical reason to believe that future game schedules will distributionally adhere to previous data, particularly given conference re-alignments and changes in television contracts, so permuting existing data would not account for all potential outcomes.

## 5. Conclusion

We need to rigorously interrogate assumptions concerning what affects performance in team sports. There is no clear indication that jet lag and game time affect team performance when appropriate analyses are performed in a causal inference framework. Similarly rigorous analyses should be undertaken to investigate other assumptions in sport science, such as workload management, sleep practices, and dietary/supplementation regimens.

## Data Availability

All data in the present study will be made publicly available upon acceptance in a peer-reviewed journal.

## References

1. Reilly T, Waterhouse J, Edwards B. Jet Lag and Air Travel: Implications for Performance. Clin Sports Med. 2005;24(2):367–380. doi:10.1016/j.csm.2004.12.004

2. Reilly T, Atkinson G, Waterhouse J. Travel fatigue and jet-lag. J Sports Sci. 1997;15(3):365-369. doi:10.1080/026404197367371

3. Forbes-Robertson S, Dudley E, Vadgama P, Cook C, Drawer S, Kilduff L. Circadian Disruption and Remedial Interventions. Sports Med. 2012;42(3):185-208. doi:10.2165/11596850-000000000-00000

4. Manfredini R, Manfredini F, Fersini C, Conconi F. Circadian rhythms, athletic performance, and jet lag. Br J Sports Med. 1998;32(2):101–106. doi:10.1136/bjsm.32.2.101

5. Lee A, Galvez JC. Jet Lag in Athletes. Sports Health. 2012;4(3):211–216. doi:10.1177/1941738112442340

6. Janse van Rensburg DC, Jansen van Rensburg A, Fowler PM, et al. Managing Travel Fatigue and Jet Lag in Athletes: A Review and Consensus Statement. Sports Med. 2021;51(10):2029–2050. doi:10.1007/s40279-021-01502-0

7. Reilly T, Atkinson G, Edwards B, et al. Coping with jet-lag: A Position Statement for the European College of Sport Science. Eur J Sport Sci. 2007;7(1):1–7. doi:10.1080/17461390701216823

8. Waterhouse J, Reilly T, Atkinson G, Edwards B. Jet lag: trends and coping strategies. The Lancet. 2007;369(9567):1117-1129. doi:10.1016/S0140-6736(07)60529-7

9. Arendt J. Managing jet lag: Some of the problems and possible new solutions. Sleep Med Rev. 2009;13(4):249–256. doi:10.1016/j.smrv.2008.07.011

10. Medicine AA of S. International classification of sleep disorders—third edition (ICSD-3). AASM Resour Libr. 2014;281:2313.

11. Fowler P, Duffield R, Vaile J. Effects of simulated domestic and international air travel on sleep, performance, and recovery for team sports. Scand J Med Sci Sports. 2015;25(3):441–451. doi:10.1111/sms.12227

12. Fowler P, Duffield R, Vaile J. Effects of Domestic Air Travel on Technical and Tactical Performance and Recovery in Soccer. Int J Sports Physiol Perform. 2014;9(3):378–386. doi:10.1123/ijspp.2013-0484

13. Richmond LK, Dawson B, Stewart G, Cormack S, Hillman DR, Eastwood PR. The effect of interstate travel on the sleep patterns and performance of elite Australian Rules footballers. J Sci Med Sport. 2007;10(4):252–258. doi:10.1016/j.jsams.2007.03.002

14. Song A, Severini T, Allada R. How jet lag impairs Major League Baseball performance. Proc Natl Acad Sci. 2017;114(6):1407–1412. doi:10.1073/pnas.1608847114

15. Leota J, Hoffman D, Czeisler MÉ, et al. Eastward Jet Lag is Associated with Impaired Performance and Game Outcome in the National Basketball Association. Front Physiol. 2022;13. Accessed August 16, 2023. https://www.frontiersin.org/articles/10.3389/fphys.2022.892681

16. Glinski J, Chandy D. Impact of jet lag on free throw shooting in the National Basketball Association. Chronobiol Int. 2022;39(7):1001–1005. doi:10.1080/07420528.2022.2057321

17. Roy J, Forest G. Greater circadian disadvantage during evening games for the National Basketball Association (NBA), National Hockey League (NHL) and National Football League (NFL) teams travelling westward. J Sleep Res. 2018;27(1):86–89. doi:10.1111/jsr.12565

18. Smith RS, Efron B, Mah CD, Malhotra A. The Impact of Circadian Misalignment on Athletic Performance in Professional Football Players. Sleep. 2013;36(12):1999–2001. doi:10.5665/sleep.3248

19. Recht LD, Lew RA, Schwartz WJ. Baseball teams beaten by jet lag. Nature. 1995;377(6550):583-583. doi:10.1038/377583a0

20. Nissen SB, Magidson T, Gross K, Bergstrom CT. Publication bias and the canonization of false facts. Rodgers P, ed. eLife. 2016;5:e21451. doi:10.7554/eLife.21451

21. Little RJ, Lewis RJ. Estimands, Estimators, and Estimates. JAMA. 2021;326(10):967-968. doi:10.1001/jama.2021.2886

22. Pearl J. Causal Analysis in Theory and Practice. Published 2019. Accessed October 28, 2022. http://causality.cs.ucla.edu/blog/index.php/page/3/

23. Candlish J, Teare MD, Dimairo M, Flight L, Mandefield L, Walters SJ. Appropriate statistical methods for analysing partially nested randomised controlled trials with continuous outcomes: a simulation study. BMC Med Res Methodol. 2018;18(1):105. doi:10.1186/s12874-018-0559-x

24. Moerbeek M, Schie S van. What are the statistical implications of treatment non-compliance in cluster randomized trials: A simulation study. Stat Med. 2019;38(26):5071–5084. doi:10.1002/sim.8351

25. Vorland CJ, Brown AW, Dawson JA, et al. Errors in the implementation, analysis, and reporting of randomization within obesity and nutrition research: a guide to their avoidance. Int J Obes. 2021;45(11):2335–2346. doi:10.1038/s41366-021-00909-z

26. Rubin DB. Estimating causal effects of treatments in randomized and nonrandomized studies. J Educ Psychol. 1974;66:688–701. doi:10.1037/h0037350

27. Pearl J. Causal Diagrams for Empirical Research. Biometrika. 1995;82(4):669–688. doi:10.2307/2337329

28. Scotina AD, Gutman R. Matching algorithms for causal inference with multiple treatments. Stat Med. 2019;38(17):3139–3167. doi:10.1002/sim.8147

29. Mayer I, Zhao P, Greifer N, Huntington-Klein N, Josse J. CRAN Task View: Causal Inference. Published August 4, 2023. Accessed November 16, 2023. https://CRAN.R-project.org/view=CausalInference

30. Schuler MS, Chu W, Coffman D. Propensity score weighting for a continuous exposure with multilevel data. Health Serv Outcomes Res Methodol. 2016;16(4):271–292. doi:10.1007/s10742-016-0157-5

31. Arpino B, Mealli F. The specification of the propensity score in multilevel observational studies. Comput Stat Data Anal. 2011;55(4):1770–1780. doi:10.1016/j.csda.2010.11.008

32. Royston P, Ambler G. GAM: Stata Module for Generalised Additive Models.; 2012. Accessed August 29, 2023. https://econpapers.repec.org/software/bocbocode/s428701.htm

33. Wood SN. Generalized Additive Models: An Introduction with R, Second Edition. 2nd ed. Chapman and Hall/CRC; 2017. doi:10.1201/9781315370279

34. Wood SN. On Confidence Intervals for Generalized Additive Models Based on Penalized Regression Splines. Aust N Z J Stat. 2006;48(4):445–464. doi:10.1111/j.1467-842X.2006.00450.x

35. Wang Yuedong, Wahba G. Bootstrap confidence intervals for smoothing splines and their comparison to bayesian confidence intervals. J Stat Comput Simul. 1995;51(2-4):263–279. doi:10.1080/00949659508811637

36. Robertson SE, Steingrimsson JA, Joyce NR, Stuart EA, Dahabreh IJ. Estimating subgroup effects in generalizability and transportability analyses. Am J Epidemiol. Published online February 28, 2022:kwac036. doi:10.1093/aje/kwac036

37. Glynn AN, Quinn KM. An Introduction to the Augmented Inverse Propensity Weighted Estimator. Polit Anal. 2010;18(1):36–56. doi:10.1093/pan/mpp036

38. Gerber AS, Green DP. Field Experiments: Design, Analysis, and Interpretation. Illustrated edition. W. W. Norton & Company; 2012.

39. Zhang Y, Zhao Q. What is a Randomization Test? J Am Stat Assoc. 2023;0(0):1–15. doi:10.1080/01621459.2023.2199814

40. Teucher A, Rudis B. Lutz: Look Up Time Zones of Point Coordinates.; 2019. Accessed August 29, 2023. https://cran.r-project.org/web/packages/lutz/index.html

41. Robinson D, Bryan J, Elias J. Fuzzyjoin: Join Tables Together on Inexact Matching.; 2020. Accessed August 29, 2023. https://cran.r-project.org/web/packages/fuzzyjoin/index.html

42. List of NCAA Division I institutions. In: Wikipedia. ; 2023. Accessed August 18, 2023. https://en.wikipedia.org/w/index.php?title=List_of_NCAA_Division_I_institutions&oldid=1170365859

43. Abatzoglou JT. Development of gridded surface meteorological data for ecological applications and modelling. Int J Climatol. 2013;33(1):121–131. doi:10.1002/joc.3413

44. Gilani S, Easwaran A, Lee J, et al. CfbfastR: Access College Football Play by Play Data.; 2022. Accessed August 29, 2023. https://cran.r-project.org/web/packages/cfbfastR/index.html

45. Attia J, Holliday E, Oldmeadow C. A proposal for capturing interaction and effect modification using DAGs. Int J Epidemiol. 2022;51(4):1047–1053. doi:10.1093/ije/dyac126

46. Brookhart MA, Schneeweiss S, Rothman KJ, Glynn RJ, Avorn J, Stürmer T. Variable selection for propensity score models. Am J Epidemiol. 2006;163(12):1149–1156. doi:10.1093/aje/kwj149

47. Fong C, Hazlett C, Imai K. Covariate balancing propensity score for a continuous treatment: Application to the efficacy of political advertisements. Ann Appl Stat. 2018;12(1):156–177.

48. Singer JD. Using SAS PROC MIXED to Fit Multilevel Models, Hierarchical Models, and Individual Growth Models. J Educ Behav Stat. 1998;23(4):323–355. doi:10.3102/10769986023004323

49. Tenan MS, Marti CN, Griffin L. Motor unit discharge rate is correlated within individuals: a case for multilevel model statistical analysis. J Electromyogr Kinesiol Off J Int Soc Electrophysiol Kinesiol. 2014;24(6):917–922. doi:10.1016/j.jelekin.2014.08.014

50. Austin PC. Assessing covariate balance when using the generalized propensity score with quantitative or continuous exposures. Stat Methods Med Res. 2019;28(5):1365–1377. doi:10.1177/0962280218756159

51. Austin PC, Stuart EA. Moving towards best practice when using inverse probability of treatment weighting (IPTW) using the propensity score to estimate causal treatment effects in observational studies. Stat Med. 2015;34(28):3661–3679. doi:10.1002/sim.6607

52. Rassen JA, Brookhart MA, Glynn RJ, Mittleman MA, Schneeweiss S. Instrumental variables II: instrumental variable application—in 25 variations, the physician prescribing preference generally was strong and reduced covariate imbalance. J Clin Epidemiol. 2009;62(12):1233–1241. doi:10.1016/j.jclinepi.2008.12.006

53. Zhu Y, Coffman DL, Ghosh D. A Boosting Algorithm for Estimating Generalized Propensity Scores with Continuous Treatments. J Causal Inference. 2015;3(1):25–40. doi:10.1515/jci-2014-0022

54. Hox J, Moerbeek M, van de Schoot R. Multilevel Analysis: Techniques and Applications, Third Edition. 3rd ed. Routledge; 2018. doi:10.4324/9781315650982

55. Lucas RM. Frequencies, Unequal Variance Weights, and Sampling Weights: Similarities and Differences in SAS. In: SAS Global Forum 2018. Vol 1938-2018. SAS; 2018:1-14. https://support.sas.com/resources/papers/proceedings18/1938-2013.pdf

56. Westreich D, Cole SR. Invited Commentary: Positivity in Practice. Am J Epidemiol. 2010;171(6):674–677. doi:10.1093/aje/kwp436

57. Chattopadhyay A, Hase CH, Zubizarreta JR. Balancing vs modeling approaches to weighting in practice. Stat Med. 2020;39(24):3227–3254. doi:10.1002/sim.8659

58. Efron B. Bootstrap Methods: Another Look at the Jackknife. Ann Stat. 1979;7(1):1–26. doi:10.1214/aos/1176344552

59. Heß S. Randomization Inference with Stata: A Guide and Software. Stata J. 2017;17(3):630–651. doi:10.1177/1536867X1701700306

60. Ernst MD. Permutation Methods: A Basis for Exact Inference. Stat Sci. 2004;19(4):676–685.

61. Lehmann EL, Romano JP. Testing Statistical Hypotheses. Springer International Publishing; 2022. doi:10.1007/978-3-030-70578-7

62. Kempthorne O, Doerfler TE. The Behaviour of Some Significance Tests Under Experimental Randomization. Biometrika. 1969;56(2):231–248. doi:10.2307/2334417

